# Fatal cases after Omicron BA.1 and BA.2 infection: Diffuse alveolar damage occurs only in a minority – results of an autopsy study

**DOI:** 10.1101/2022.10.02.22280609

**Authors:** Bruno Märkl, Sebastian Dintner, Tina Schaller, Eva Sipos, Elisabeth Kling, Silvia Miller, Francisco Farfan, Przemyslaw Grochowski, Nic Reitsam, Johanna Waidhauser, Klaus Hirschbühl, Oliver Spring, Andre Fuchs, Thomas Wibmer, Peter Boor, Martin Beer, Claudia Wylezich

## Abstract

Compared with previously prevalent variants of SARS-CoV-2, the Omicron lineages BA.1 and BA.2 are known to be associated with mild clinical courses. In addition, well-established animal models do not develop severe diseases.

To address whether the supposedly fatal cases after Omicron-BA.1/2 infection show the known COVID-19 organ alterations, especially in the lungs, 23 full and 3 partial autopsies in the deceased with known Omicron BA.1/2 infections have been consecutively performed. Viral RNA was determined by RT-qPCR and RNA-in situ hybridization. The lineages were analyzed by whole genome sequencing or S-gene analysis.

Despite high viral loads in almost all nasopharyngeal swabs and in 13 lung tissue samples, death caused by COVID-19-associated diffuse alveolar damage (DAD) in the acute and organizing stages was found in only eight cases (31%). This rate is significantly lower compared to previous studies, including non-Omicron variants, where rates of 92% and 69% for non-vaccinated and fully vaccinated vaccines were observed. It is of special interest that neither vaccination status nor known risk factors (i.e., age, comorbidities, obesity, immuno-suppression) were significantly associated with a direct cause of death by COVID-19. Only the reason for the hospital admission of the patients due to COVID-19-related symptoms showed a significant correlation with directly COVID-19-caused deaths (P < 0.001).

DAD still occurred in the Omicron BA.1/BA.2 era of the SARS-CoV-2 pandemic but at a considerably lower frequency than seen with previous variants of concern. In our study, none of the known risk factors discriminated the cases with COVID-19-caused death from those that had COVID-19 infections but died due to a different disease. Therefore, the host’s genomics might play a key role in this regard. Further studies are urgently needed to elucidate the existence of a genomic mechanism as a risk factor for a fatal course.

## Introduction

The Omicron variant of concern (VOC) of SARS-CoV-2 is characterized by both high infectivity and transmissibility. Despite this, it causes a rather mild clinical course of COVID-19 compared to the other VOCs (1-3). In concordance, animal experiments showed reduced pathogenicity of the Omicron variants BA1 and BA2 compared to other VOCs. This includes less prominent loss of weight and lower viral burden in the upper and lower respiratory tracts in hamsters, ACE2-wildtype mice, and K18-hACE2 transgenic mice (4). In hamsters, the Delta variant was dominant over the BA1 lineage of Omicron. In ferrets, COVID-19 infection was even abortive (5). Meanwhile, five lineages (BA.1, 2, 3, 4, and 5) have been identified and characterized (6). Because the Omicron variant is associated with a significant immune escape, both the effectiveness of current vaccines and the immunity of convalescents are hampered (7-10). Despite this immune escape, booster vaccination has also been reported to reduce the mortality of COVID-19 caused by the Omicron variant (11). Although a considerable number of studies regarding the Wuhan strain and non-Omicron variants report effects on nearly all human organ systems, the direct cause of death is COVID-19 pneumonia with different stages of diffuse alveolar damage (DAD) (12, 13). The involvement of the vasculature seems to be responsible, in part, for these severe lung injuries (14). A dysfunctional immune reaction after SARS-CoV-2 infection that causes an immediate release of cytokines and a self-amplifying mechanism leading to a cytokine storm is likely causative of the COVID-19 involvement of many organs without direct viral interaction (15). Given the data on reduced pathogenicity of the Omicron variant compared to previous VOCs, the question arises of whether and to what extent COVID-19 pneumonia is prevalent and of the cause of death in the deceased with Omicron SARS-CoV-2-infection. To address this question, we analyzed the cases of the Augsburg autopsy study with confirmed Omicron variant BA1/2 infections.

## Material and Methods

### Case collection

The Omicron study cohort comprises 26 deceased with proven infection with one of the known Omicron lineages of SARS-CoV-2 between January 2022 and May 2022. All patients were treated at the University Medical Center of Augsburg. In total, 160 autopsy cases of infections of other SARS-CoV-2 variants, partially included in previously published studies, served as controls (12, 16, 17). Full autopsies were performed in 23 cases, while in three cases, the relatives restricted the autopsy to a minimally invasive approach.

Informed consent was obtained from the next of kin. The study was approved by the ethics committee of the Ludwig Maximilian University Munich (Project numbers 20–426; 22-0469)

### Autopsy, sample collection, histology

The techniques and procedures used for the autopsy, sample collection, and histology have been described previously (16). In brief, autopsies were performed within a body bag with respect to adequate safety rules (18, 19). Full autopsies included the opening of all body cavities and careful inspection and tissue sampling of all organs. In partial autopsies, larger tissue samples from the thoracal and abdominal organs were obtained from epigastric access. Regarding the causes of death, we classified the diagnosis that led directly to death. Importantly, this is different from the WHO definition, which includes COVID-19 as a cause of death in cases when an existing disease is exacerbated due to COVID-19 (20). Histological analyses are based on hematoxylin & eosin (HE) and periodic acid-Schiff stains (PAS). No immunohistochemical staining was performed.

### RNA-in situ hybridization

These techniques have also been described previously (17). In brief, RNA-in situ hybridization (ISH) was performed on representative lung samples from all Omicron cases using SARS-CoV-2 RNA-specific antisense probes designed and synthesized by Advanced Cell Diagnostics (ACD, Palo Alto, CA, USA; Cat. No: 848568). The RNAscope ISH assays were conducted on the Leica BOND-RX System (Leica, Germany) using the RNAscope 2.5 LS Reagent kit-BROWN (ACD, Cat. No: 322100). Chromogen detection and hematoxylin counterstaining were performed using a bond polymer refine detection kit (Leica, Cat. No.: DS9800).

### RT-qPCR

The RT-qPCR assay was described earlier. Briefly, RNA was extracted from FFPE sections using the Maxwell CSC RNA FFPE Kit (AS1360, Promega) and swabs using the Maxwell 16 LEV Blood DNA Kit (AS1290, Promega) on a Maxwell system (Promega Corporation, Madison, WI, USA). An MS2 phage control was added to the samples before the extraction of the RNA. A negative control containing only MS2 Phage was prepared and used for RT-qPCR after extraction. RT-qPCR was performed on a QuantStudio 5 Dx real-time PCR instrument (Thermo Fisher, Carlsbad, CA, USA) using the Taq-Path COVID-19 CE-IVD RT-PCR Kit (Thermo Fisher, Pleasanton, TX, USA). The cycle threshold (Ct) values were classified in six categories (<10; 11–17; 18–24; 25–29; 30–40; negative). In cases where whole-genome sequencing of the virus was not available, the Omicron BA.1 lineage was determined by S-Gene target failure (STGF). S-Gene negative cases were assigned to the BA.1 group. Viral dissemination with widespread viral RNA detection was defined in (17).

### Viral whole genome sequencing/variant determination

SARS-CoV-2 whole-genome sequences were generated using a generic metagenomics workflow (21). This procedure was combined with a capture enrichment procedure using SARS-CoV-2 specific myBaits (Daicel Arbor Biosciences, Ann Arbor, USA), if necessary (22). In some cases, the Ion AmpliSeq SARS- CoV- 2 Research Panel (Thermo Fisher Scientific, Germany) was applied using the Ion Chef instrument. After quality checks and quantification of generated sequencing libraries, they were pooled together and sequenced on an Ion Torrent S5XL instrument (Thermo Fisher Scientific, Germany) with Ion 530 sequencing chips and chemistry for 400 base pair reads. Raw sequencing data were analyzed using the Genome Sequencer Software Suite (version 2.6; Roche, Mannheim, Germany https://roche.com), with default software settings for quality filtering and mapping, using the SARS-CoV-2 reference sequence Wuhan-Hu-1 (MN908947). SARS-CoV-2 lineages were determined with the Pangolin COVID-19 Lineage Assigner (23). In addition, the obtained SARS-CoV-2 genome sequences were aligned together with sequences retrieved from GenBank and Gisaid using MAFFT version 7.38837, as implemented in Geneious version 10.2.3 (Biomatters, Auckland, New Zealand). Phylogenetic trees were constructed with PhyML version 3.038, using the GTR+ GAMMA+ I model with 100 bootstrap replications, and MrBayes version 3.2.639, using the GTR model with eight rate categories and a proportion of invariable sites in the Geneious software package. The Bayesian analysis was performed for 1,000,000 generations and sampled every 1,000 generations for four simultaneous chains. The SARS-CoV-2 genome sequences generated in this study are available under accession numbers OP430881–OP430898.

### Statistics

Categorial data were compared using the Chi-square or Fisher’s exact test depending on the group sizes. Depending on the distribution status and group numbers, the student’s-test, the Mann-Whitney Rank sum test, or analysis of variance (ANOVA) were used to compare continuous data. Correlations between non-constant variables were calculated using Spearman rank order correlation. A p-value of less than 0.05 was considered significant. All analyses were calculated using the Sigma Plot 13.0 software package (Systat, San Jose, CA, USA).

## Results

### Study cohort, vaccination status, causes of death

During the study period, 138 patients died from a proven SARS-CoV-2 infection at the University Medical Center in Augsburg. The study cohort comprises 26 consecutively collected cases, resulting in an autopsy rate of 19%. The demographic and clinicopathological data are summarized in Table 1. Six (23%) of the deceased were non-vaccinated, three (12%) were partially vaccinated, and 17 patients were fully vaccinated (65%), including 10 (38%) cases with one and two (8%) with two booster vaccinations. The rate of fully vaccinated persons among the inhabitants of the city of Augsburg was 77% (July 2022) (24), which means that the group of fully vaccinated persons is slightly underrepresented in this study but without statistical significance (P = 0.253).

**Table 1:** Clinicopathological Data.

|  | Entire Omicron-Study-Group<br>n = 26 | Non-COVID-19 death<br>n = 18 | COVID-19 death<br>n = 8 | P-Value |
| --- | --- | --- | --- | --- |
| Median Age [years] | 82 (52 - 92) | 83 (52 - 92) | 80 (58 - 90) | 0.656 |
| Sex [f : m] | 1 : 3 | 1 : 2 | 0 : 8 | 0.132 |
| Median number of comorbidities | 4 (0 - 7) | 4 (0 - 7) | 4 (0 - 5) | 0.683 |
| Cancer | 8 (31%) | 5 (28%) | 3 (38%) | 0.667 |
| COVID-19 hospital admission* | 8 (31%) | 1 (6%) | 7 (88%) | < 0.001 |
| Median BMI [kg/m<sup>2</sup>] | 28 (16 - 48) | 28 (16 - 48) | 29 (19 - 33) | 0.662 |
| Hint for Immuno suppression | 11 (42%) | 7 (39%) | 4 (50%) | 0.683 |
| Invasive Ventilation | 6 (23%) | 3 (17%) | 3 (38%) | 0.33 |
| Omicron Variant BA. 1 | 21 (81%) | 15 (83%) | 6 (75%) |  |
| Omicron Variant BA. 2 | 5 (19%) | 3 (17%) | 2 (25%) | 0.628 |
| non-vaccinated | 6 (23%) | 5 (28%) | 1 (13%) |  |
| partial vaccinated | 3 (12%) | 3 (17%) | 0 |  |
| twice vaccinated | 5 (19%) | 3 (17%) | 2 (25%) | 0.190<sup>#</sup> |
| one booster vaccination | 10 (38%) | 6 (33%) | 4 (50%) |  |
| two booster vaccinations | 2 (8%) | 1 (6%) | 1 (13%) |  |
| Median Ct-value nasophar. (initial) | 27 (24 - 38) | 27 (24 - 38) | 27 (24 - 29) | 0.903 |
| Median Ct-value nasophar. (Autopsy) | 20 (10 - 34) | 22 (10 - 34) | 19 (13 - 30) | 0.594 |
| IgA-levels (normal: 70 – 400)[mg/dl] | 241 (14 - 453) | 402 (166 - 453) | 212 (14 - 426) | 0.077 |
| IgG-levels (normal: 700 – 1600)[mg/dl] | 759 (286 - 1360) | 1185 (286 - 1340) | 692 (491 - 1360) | 0.338 |
| Highest CRP (normal: <0.5)[mg/dl] | 11 (0.8 - 43) | 10 (0.8 - 43.0) | 12.0 (1.7 - 27.4) | 0.598 |
| Highest procalcitonin (normal: <0.5)[ng/ml] | 0.9 (0.1 - 55) | 0.4 (0.1 - 55) | 1.2 (0.1 - 25.0) | 0.535 |
| Highest IL-6 (normal: <15)[pg/ml] | 500 (17 - 3560) | 392 (17 - 3560) | 728 (31 - 2570) | 0.620 |
| Time from first symptom to death [days] | 9 (1 - 44) | 9 (1 - 23) | 17 (5 - 44) | 0.096 |
| Time from first positive PCR to death [days] | 7 (1 - 51) | 5 (1-51) | 17 (1 -44) | 0.094 |
| **Non COVID-19 direct causes of death:** |  |  |  |  |
| Acute pneumonia |  | n.a. | 5 |  |
| Cardiac failure |  | n.a. | 7 |  |
| Probably arrhythmia |  | n.a. | 1 |  |
| Mesenterial infarction |  | n.a. | 1 |  |
| Pulmonary embolism |  | n.a. | 1 |  |
| Myocardial infarction |  | n.a. | 1 |  |
| Sepsis |  | n.a. | 2 |  |

\* Number of patients with hospital admission due primarily to COVID-19; # P-value was calculated between un-/incomplete vaccinated versus at least two vaccinations or booster received.

Based on the autopsy results, in the group of un- or partially vaccinated patients, only one person died directly due to COVID-19 pneumonia (11%), compared to seven (41%) (P = 0.190) in the group of vaccinated patients, resulting in a total rate of 31% COVID-19-caused deaths. The corresponding rates in previous studies that served as controls were 92% and 69% for non-vaccinated and fully vaccinated patients, respectively (17). The non-COVID-19 causes of death are summarized in Table 1. In 13 patients who administratively belonged to the Public Health Department of the city of Augsburg, the cause of death provided to the official disease surveillance system was determined. Compared to the autopsy results, one case was consistently classified as caused by COVID-19. In eight cases, COVID-19 was classified divergently. In four cases, death was stated not to be due to COVID-19 in accordance with the autopsy results.

### Organ involvement: Histology, RNA-ISH, PCR

The histological evaluation of all lung tissue samples revealed severe DAD of different stages in eight (31%) cases (Figure 1A). In those cases, consecutive respiratory failure was the direct cause of death. In two additional cases, a mild non-fatal DAD was identified but did not directly cause the deaths. Severe acute pneumonia with dense infiltration of one or even two entire lobes was found in eight cases (31%) (Figure 1D). Overlap with severe, partly organizing DAD occurred in only one of those acute pneumonia cases (Figure 1B). Aspergillosis was also identified in one of these cases. In concordance with previous investigations (16), no other organs showed alterations that could be classified as SARS-CoV-2-specific on the level of conventional light microscopy. SARS-CoV-2 identification by RNA-ISH was performed in samples from all lungs and revealed positive results in ten cases, with a highly significant correlation with the results of the RT-PCR (P < 0.001) from the lung samples (Figure 2B). The median Ct-value of the nasopharyngeal RT-PCR was 20 (range: 10–43). In five cases (19%), viral dissemination within the organ system, as previously defined and reported (17), is in the same range as in our series of non-VOC (16) but significantly lower than in our previous analyses of vaccinated non-Omicron cases (P = 0.014). Only one of these five cases belongs to the group of COVID deaths.

**Figure 1:**
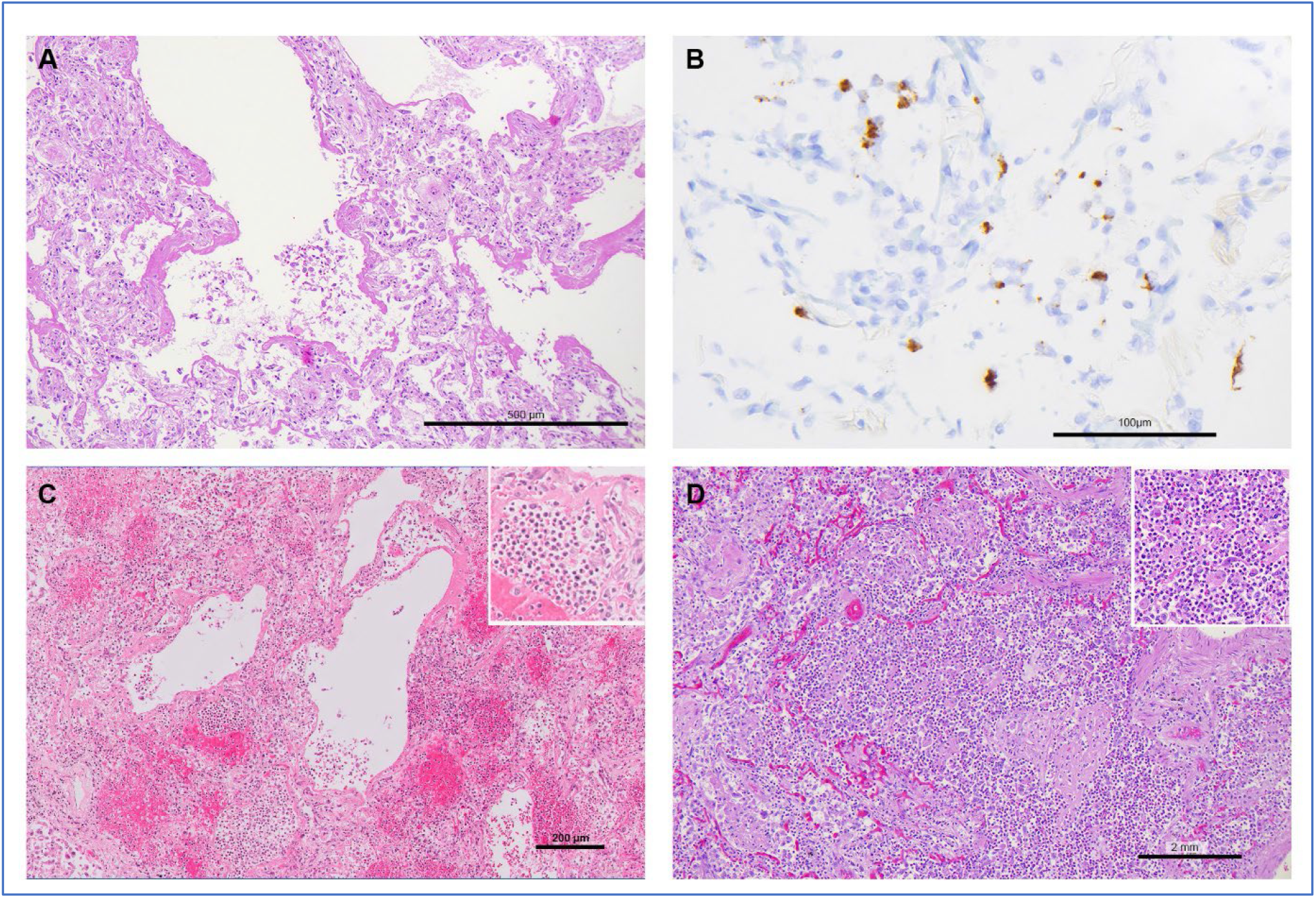
A) H&E stain; acute DAD with alveolar spaces aligned with hyaline membrane. B) SARS-CoV-2 RNA in-situ hybridization; detection of viral RNA within the lung parenchymal. C) H&E stain; simultaneous development of DAD and acute pneumonia with dense neutrophilic infiltration. Insert: higher magnification, same case. D) H&E stain; severe acute pneumonia with parenchymal destruction and dense neutrophilic infiltration. Insert: higher magnification, same case.

**Figure 2:**
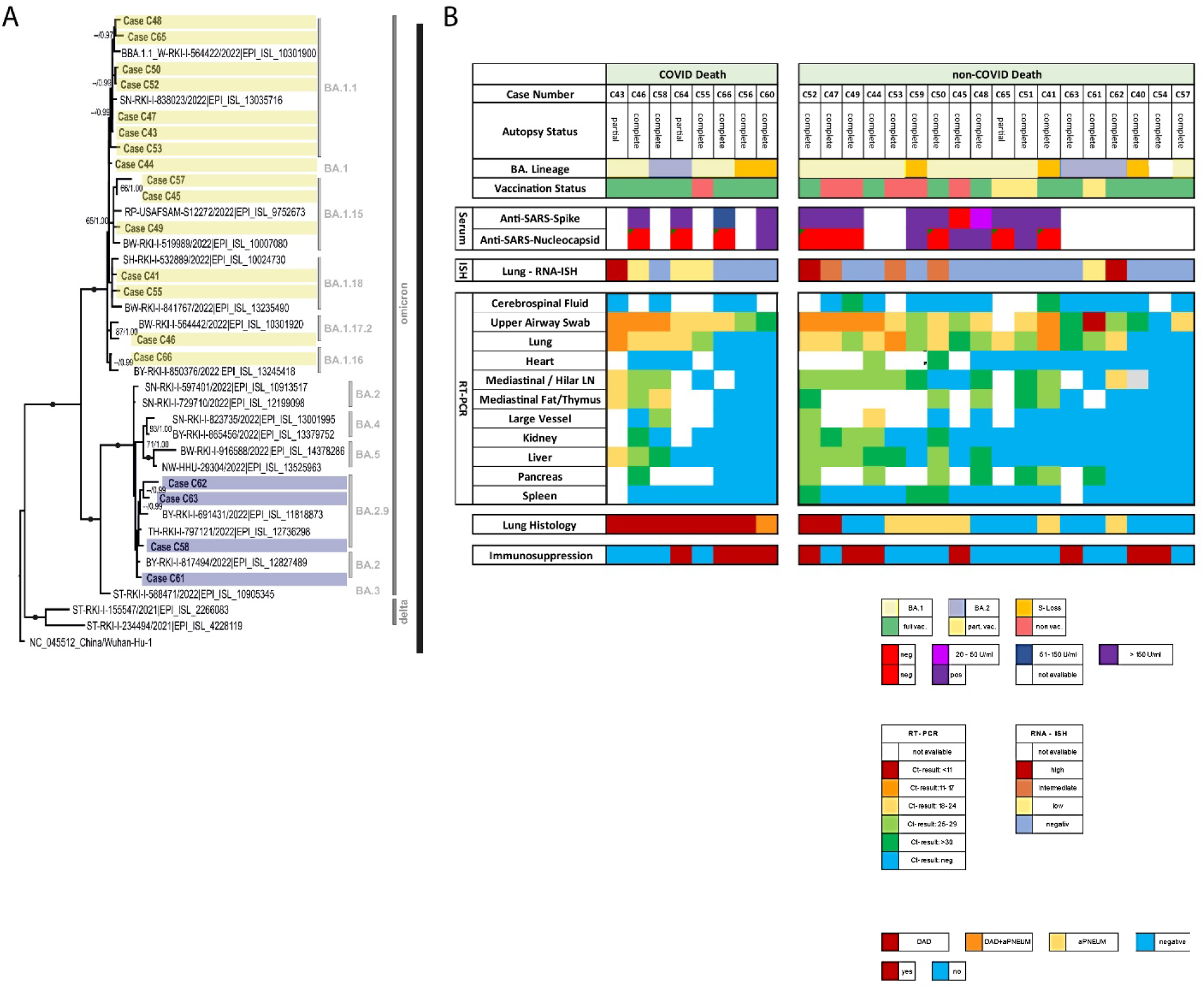
**A)** Phylogenetic tree representing cases in which the whole viral genome could be sequenced. Dots indicate bootstrap values of 100/1.00 (Maximum Likelihood/MrBayes). Support values above 50% are given. B) Autopsy-status, viral variant lineages, anti-SARS-antibody titer, and viral infection in different organs by RT-qPCR and RNA-ISH (for lungs only). S-Loss = S-Gene loss determined by RT-qPCRPCR; aPNEUM = severe acute pneumonia; hint for immunosuppression includes low immune globulin level, cancer, and drug-related; COVID-19 Death = cause of death is directly COVID-19 related. Note: this is not identical with the WHO definition. Lung histology indicates the occurrence of DAD and acute pneumonia. In cases C52 and C47, DAD was mild without respiratory impairment. Note: Cases are not sorted in a consecutive manner but by grade of viral dissemination.

### Serum analyses and sequencing

In 14 cases, the serum anti-SARS-spike and the anti-SARS-nucleocapsid antibodies were evaluated (Figure 2B). In all except one case, anti-SARS-spike antibodies were identified. This was especially true in all investigated fully vaccinated cases. In only five cases, anti-nucleocapsid antibodies were found, and only two of these belonged to the fully vaccinated group. Other serum values, including IL6, are summarized in Table 1. All infections could be assigned either to the SARS-CoV-2 Omicron BA.1 (n = 21) or BA.2 (n = 5) lineages. Detailed assignment to sub-lineages was possible for cases that provided a full or nearly full genome sequence (Figure 2A). In one case (C54), no RNA was available, but this case occurred when BA.1 sublineages were circulating. None of the clinic-pathological or outcome parameters correlated with the attributed Omicron lineage.

### Comparison between cases with and without direct COVID-19-related deaths

The comparison between cases with and without direct COVID-19-related deaths revealed no discriminating factor except the clinical presentation (Table 1). Seven out of eight (88%) fatal cases presented as COVID-19 on admission, while the admission in only 6% of non-fatal cases was directly COVID-19 related (P < 0.001). The vaccination status did not significantly differ between the groups. In the 94% of remaining cases, the hospital admission was primarily due to other diseases. There was a clear trend toward a more prolonged period between first symptoms/positive PCR testing and death in COVID-19-caused deaths compared to other causes of death (P = 0.096 and 0.094). Immune globulin levels tended to be decreased in COVID-19 deaths, with a statistical trend for IgA (P = 0.077). All other investigated clinicopathological data, including age, sex, BMI, comorbidities, and cancer history, hint at immune suppression and did not differ between the two groups (Table 1).

## Discussion

To the best of our knowledge, we present the results of the first autopsy study of the deceased with proven SARS-CoV-2 Omicron BA.1 or BA.2 infection. This study delivers three main findings.

First, the frequency of directly COVID-19-caused deaths decreased considerably from initially 92% (non-Omicron, non-vaccinated COVID-19 deceased (16)) to 72% (non-Omicron, partially or fully vaccinated COVID-19 deceased, (17)) to 31% in the current study (Omicron). Second, although it is a rare scenario, severe fatal COVID-19 caused by Omicron BA.1/BA.2 does not differ regarding organ involvement and morphology compared to previous VOCs and the Wuhan strain. Third, no convincing factor could be identified to discriminate between patients who were prone to develop severe fatal COVID-19 and the majority of individuals who died with SARS-CoV-2 infection but due to a different disease.

There is a large body of evidence that infections with the SARS-CoV-2 Omicron lineages differ in several terms from those of previous prevalent strains and VOCs (1, 3). These differences concern increased infectivity combined with a relevant immune escape of Omicron strains (Ref). Infections are related to a lower risk for hospitalization and severe clinical courses, including death (1, 3, 25-28). This is in concordance with our result of a significantly reduced rate of direct COVID-19-caused deaths. Additionally, we identified a high rate of breakthrough infections in 65% of all cases. Twelve of those 17 cases received at least one booster vaccination. In addition, seven out of eight fatal COVID-19 courses were fully vaccinated, and two had even received the second booster. This underlines the ability of the Omicron lineages to escape efficiently from ancestral vaccine-derived immunity. However, at this point, it must be emphasized that our study is limited by a small number of cases. This is also because of an autopsy rate of only 19%, which is considerably decreased compared to previous studies, with 86% (16) and 55% (17), respectively. Large population-based investigations report reduced effectiveness of vaccines in preventing fatal course. However, a relevant protective effect is still reported (1, 28, 29). The results of this study allow for no direct transfer to the general population. However, they suggest that in the era of Omicron BA.1/BA.2, some patients with the Omicron variant still die due to classical COVID-19 pneumonia that shows the same morphology as with previous SARS-CoV-2 variants. This is an important finding because, as discussed above, the Omicron variant is known to be attenuated regarding its potential to cause severe illness. Animal experiments using BA.1 and BA.2 strains show exclusively mild courses with almost no symptoms (4, 5).Therefore, it could be questioned whether people infected with an Omicron variant die directly from the consequences of COVID-19 at all.

The low rate of directly COVID-19-caused death may influence the official statistics of disease control institutions like the German Robert Koch-Institut. In our exemplary analysis, based on notifications according to the German infection control law (IfSG), we identified 8 out of 13 cases that were officially recorded as COVID-19-caused deaths, divergently from the results of autopsies; this may indicate a systematic problem. The establishment of regions with officially mandated post mortal investigations that result in autopsy rates that allow epidemiologic analyses could be an approach to solving this problem. The City of Hamburg followed such an approach during the first wave of the pandemic in Germany (30), but no such program currently exists in Germany. Another potential approach is to gather these data on a national level, for example, in registries such as the German Registry of COVID-19 autopsies (www.DeRegCOVID.ukaachen.de).

It would be interesting to identify factors associated with fatal SARS-COv-2 infections. However, only the reason for hospital admission and the hospital stay time differ between the groups of cases with and without death due to COVID-19. All other relevant parameters, including factors related to immune suppression, showed similar characteristics. Even remarkably low Ct-values of the upper airway swabs and the lung tissue samples indicating a high viral load occurred in cases without the development of DAD. It seems that fatal courses of BA.1/2 infections are triggered by a factor or a combination of factors that are different from those of previous SARS-CoV-2 variants, which were clearly connected with age, comorbidities, obesity, immune suppression, and vaccination status (17, 31). Increased susceptibility could be caused by the genetics of the host. For influenza

A, several genetic variations can be identified that cause inborn errors of immunity. These affect viral replication and the inflammatory response in different parts of the immune system (32, 33). Genetic variations can influence the manifestation of many infections at different levels, such as viral load, organ involvement, chronicity, malignant transformation, and severity of the acute disease (34). Very recently, a meta-analysis addressed the aspect of individual genetic susceptibility to COVID-19. Variants of four genes could be observed that are associated with an increased rate of infection, as well as variants in five genes that heighten the risk of a severe clinical course. These genes code for the angiotensin-converting enzyme, the angiotensin-II receptor 1, and tumor necrosis factor-alpha. The question of whether the different viral variants are affected similarly remains unanswered (35).

A high rate (31%) of unusually severe acute pneumonia affecting one or two entire lobes was an unexpected observation in this study. Fungal co-infections in seven out of eight (88%) autopsy cases of those deceased after long-term treatment of COVID-19 were previously described, with pulmonary involvement in four (50%) cases (36). In our previously published autopsy series of deceased vaccinated patients, we identified four out of twenty-nine (14%) cases with pulmonary aspergillosis, while in our current series, fungal infection was observed in only one (4%) case. The reported prevalence numbers of bacterial co-/superinfections in SARS-CoV-2 infections differ considerably between studies and range from less than 1% to more than 50%, depending on the clinical setting and the definition (37). There are several hypotheses about how SARS-CoV-2 infections could heighten the risk of bacterial or fungal superinfections. Epithelial disruption could enhance viral adherence, but an even more significant effect might be caused by immunogenic reactions, such as reduction of type I and III interferons, which are believed to be counterproductive regarding the defense against bacterial infections (38, 39). The question of whether the high rate of severe acute pneumonia is just an incidental accumulation, or indeed an enhanced susceptibility to superinfections, cannot be answered by this study. However, these data, together with divergent results reported in the literature, underline the need to address this issue in a separate study.

In conclusion, this study confirms that classic COVID-19 pneumonia can be caused by Omicron BA1/2 infections but at a considerably lower rate compared to previously prevalent variants. The underlying mechanism that leads to such a fatal clinical course remains unclear and needs to be elucidated in further studies, potentially including analysis of the host’s genome. Low rates of fatal courses might also influence the correctness of national statistics. The establishment of model regions with monitoring of occurring variants, patients, and inhabitants, including biobanking and performing autopsies at high frequency to be embedded in a registry such as the German COVID-19 autopsy registry (40), could be an appropriate approach to gain insight rapidly into an ongoing pandemic.

## Data Availability

All data produced in the present study are available upon reasonable request to the authors

## Acknowledgments

The authors are thankful to Kerstin Bauer, Alexandra Martin, Melanie Spörel, Christian Beul, and Patrick Zitzow for their thorough archive work and excellent technical assistance. The authors are particularly grateful to all relatives who consented to postmortem examination and thus made an invaluable contribution to the research on this new disease.

## Funding

This work was supported by the Bavarian State Ministry of Science and Arts, the German Registry of COVID-19 Autopsies (www.DeRegCOVID.ukaachen.de) and funded by the Federal Ministry of Health (ZMVI1-2520COR201), the Federal Ministry of Education and Research within the framework of the network of university medicine (NATON, 01KX2121), and the German Federal Ministry of Food and Agriculture through the Federal Office for Agriculture and Food, project ZooSeq, grant number 2819114019.

## Data Availability

Due to privacy reasons the raw data are not available on public repository. However, data in restricted form available from the corresponding authors

## Competing interests

The authors declare no competing interests.

## Notes

### Competing Interest Statement

The authors have declared no competing interest.

### Author Declarations

Ethics committee of the Ludwig Maximilian University Munich (Project numbers 20 426; 22 0469)

